# The impact of the UK’s first COVID-19 lockdown on rates of violence and aggression on psychiatric inpatient wards

**DOI:** 10.1101/2021.03.10.21253244

**Authors:** James Payne-Gill, Corin Whitfield, Alison Beck

## Abstract

**Aims:** Inpatient life in UK mental health hospitals was profoundly altered during the first wave of the COVID-19 pandemic. We analysed whether these changes impacted the rate of violent and aggressive incidents across acute adult wards and psychiatric intensive care units in a South London NHS Mental Health Trust during the first UK lockdown.

**Methods:** We used an interrupted time series analysis to assess whether the rate of violent and aggressive incidents changed during the lockdown period from 23rd March 2020 to 15th June 2020. We used a quasi-poisson general additive model to model the weekly rate of violent incidents as a function of a seasonal trend, time trend, and impact of lockdown, using data from 1^st^ January 2017 to 27^th^ September 2020.

**Results:** There was a 35% increase in the rate of incidents of violence and aggression [IR = 1.35, 95% CI: 1.15 – 1.58, p < 0.001] between March 23rd 2020 and June 15th 2020. In addition, there was strong evidence of temporal (p < 0.001) and seasonal trends (p < 0.001).

**Conclusions:** Our results suggest that restrictions to life increased the rate of violent incidents on the mental health wards studied here.

## Introduction

The SARS-CoV-2 pandemic and government-enforced lockdown limited personal freedoms, with reduced ability to travel, work, and visit friends and family. As the NHS grappled with the pandemic, changes to mental health service provision meant that some of the most vulnerable in society, psychiatric inpatients, also experienced restrictions to their daily routines and changes in their care.

Royal College of Psychiatrists, NHS England (NHSE, 2020), and Royal College of Nursing guidelines recommended a range of measures to prevent the spread of Covid-19. These included stopping ward activities that brought people into close contact, which often meant changes to ward groups, group mealtimes and visits (RCP, 2020). Given that COVID-19 attacks the lungs, guidance outlined the need to cease escorting patients out for smoking breaks. On April 8^th^, the NHS suspended hospital visits until further notice, which was not lifted until 5^th^ June when visitation procedures were left to the discretion of healthcare providers (NHSE, 2020). Access to psychological and occupational therapy was more limited, since “provision of specialist services such as occupational therapy, psychology or pharmacology is secondary to maintaining… physical health in the present situation” (RCP, 2020). NHS England also outlined the need to redeploy staff as required to meet service needs, which meant ward teams could change at short notice (NHSE, 2020). In line with advice to the public, the guidance from NHS England has been to quarantine patients who exhibit symptoms of Covid-19 in their rooms (NHSE, 2020).

While these restrictions may have been justified to protect patients, collectively they stripped freedoms away from people already deprived of many liberties. Stress on wards increased as staff and service users alike adjusted to the continued uncertainties of the virus. These factors may have had an impact on the rate of violence on wards. Before the pandemic, high rates of violence were often associated with some psychiatric admissions. Results from a meta-analysis suggest that nearly one in five inpatients commit at least one act of physical violence (Lozzino et al., 2015). One in three NHS mental health nurses reported experiencing physical violence (RCN, 2017).

A range of factors are associated with an increased risk of violence. Poor patient-staff relationships and high anxiety amongst staff have been found to increase the risk of patient violence (d’Ettorre & Pellicani, 2017). A systematic review found that 25% of all antecedents to violent incidents involve staff-patient interactions that limit patient freedoms, such as denying a patient request (Papadopoulos et al., 2012). It is therefore reasonable to assume that enforcing new restrictions, particularly around leave and smoking, may directly lead to confrontations that end in violence. A study of patients isolated in general hospitals for infection control reported higher rates depression, anxiety, and anger, as well as less time spent with healthcare workers (Abad, Fearday & Safdar, 2010). The emotional impact of being quarantined on psychiatric inpatient wards may be even more severe and increase the risk of violent confrontations. The risk of violence will also increase if the ability of staff to build therapeutic relationships is undermined. A qualitative study of service user experiences of psychiatric hospital admissions in London found that coercion undermined the building of positive relationships with staff (Gilburt, Rose & Slade, 2008). Furthermore, staff working on mental health inpatient wards find clearly defined roles, supportive ward managers, and clear organisational structures are essential to morale (Totman et al., 2011). Changes to team and organisational practices in response to the pandemic may damage staff morale, making it more difficult to manage stressful situations.

To assess whether unprecedented changes impacted rates of violence, we analysed changes in the number of incidents of violence and aggression recorded in the Trust’s incident reporting system following the release of government Covid-19 guidance for NHS mental health services.

## Methods

### Data Extraction

We extracted data on violence and aggression from the Trust’s incident reporting system, Datix. We extracted all incidents for the period 1^st^ January 2017 to 27^th^ September 2020 that involved patients on one of the Trust’s adult acute wards and Psychiatric Intensive Care Units (PICU). We included all incidents of violence and aggression because this captures a broad range of behaviours which may have been affected by the Covid-related changes to the ward environment, ranging from actual to threatened physical assault, verbal assault, challenging behaviours, and damage to property. We also extracted data on the number of occupied bed days from the Trust’s Online Analytical Processing (OLAP) cube, which extracts and aggregates data from the Trust’s electronic patient record system, ePJS, every 24 hours.

### Statistical Analysis

To assess whether changes implemented on wards had an impact on levels of violence and aggression, we conducted an interrupted time series regression analysis. We employed a similar methodology to a previous study which analysed the impact of a no-smoking policy on rates of violence in inpatients settings in the same Trust (Robson et al., 2017). We modelled the effect as a temporary level change, with the exposure beginning on 23^rd^ March 2020, when the nationwide lockdown began, and ending on 15^th^ June 2020 when the 3^rd^ and furthest easing of lockdown restrictions began. We used a generalised additive model to model the rate of weekly violent incidents as a function of an underlying time trend, seasonal trend, a binary exposure variable equal to one during the exposure period and zero otherwise, and an offset variable to account for variation in the number of patient occupied bed days. The underlying time trend was modelled using a thin plate spline, while the seasonal trend was modelled using a cyclic cubic regression spline.

## Results

Over the period 1^st^ January 2017 to 28^th^ September 2020, there was a total of 6646 incidents of violence and aggression involving patients across the Trust’s adult acute wards and Psychiatric Intensive Care Units (PICU). 1314 of these occurred in 2017, 1795 occurred in 2018, 2041 occurred in 2019, and 1496 incidents had occurred by 27^th^ September 2020. 61% of these incidents were categorised on Datix as an assault by the patient, 32% were classified as challenging behaviours, 3% were classified as damage to property, 2% were classified as harassment, and the remaining 1% of incidents were classified as sexual assaults.

Table 1 shows the results of the generalised additive model. Our analysis suggests that after adjusting for seasonal and temporal trends there is very strong evidence of an increase in the rate of violent and aggressive incidents following the Covid-19 policy changes. Our model estimates there was a 35% increase in the rate of incidents of violence and aggression [IR = 1.35, 95% CI: 1.15 – 1.58, p < 0.001] between March 23^rd^ 2020 and June 15^th^ 2020. In addition, there was strong evidence of temporal (p < 0.001) and seasonal trends (p < 0.001).

**Table 1.** Results of generalised additive model regression analysis

|  | IRR (95% CI) | p |
| --- | --- | --- |
| Covid indicator | 1.35 (1.15 – 1.58) | < 0.001 |
| Trend Terms | EDF | p |
| EDF: s(Month) | 4.70 (10.00) | < 0.001 |
| EDF: s(Time) | 2.47 (3.07) | < 0.001 |
| Deviance | 373.18 |  |
| Deviance explained | 0.35 |  |
| Dispersion | 2.21 |  |
| R<sup>2</sup> | 0.35 |  |
| GCV score | 171.32 |  |
| Num. obs. | 180 |  |
| Num. smooth terms | 2 |  |
| IRR = Incident Rate Ratio; EDF = Estimated Degrees of Freedom; GCV score = Generalised Cross-Validation score |  |  |

Figure 1 plots the weekly rate of incidents of violence or aggression per 1000 patient bed days. The red line plots the predicted rate of violence based in the full model with the 95% confidence intervals shaded out either side. The light blue line shows the predicted rate based only on the temporal and seasonal trends. There is a sharp upward jump around March 23^rd^ and a reduction leading up the 15^th^ June.

**Figure 1.**
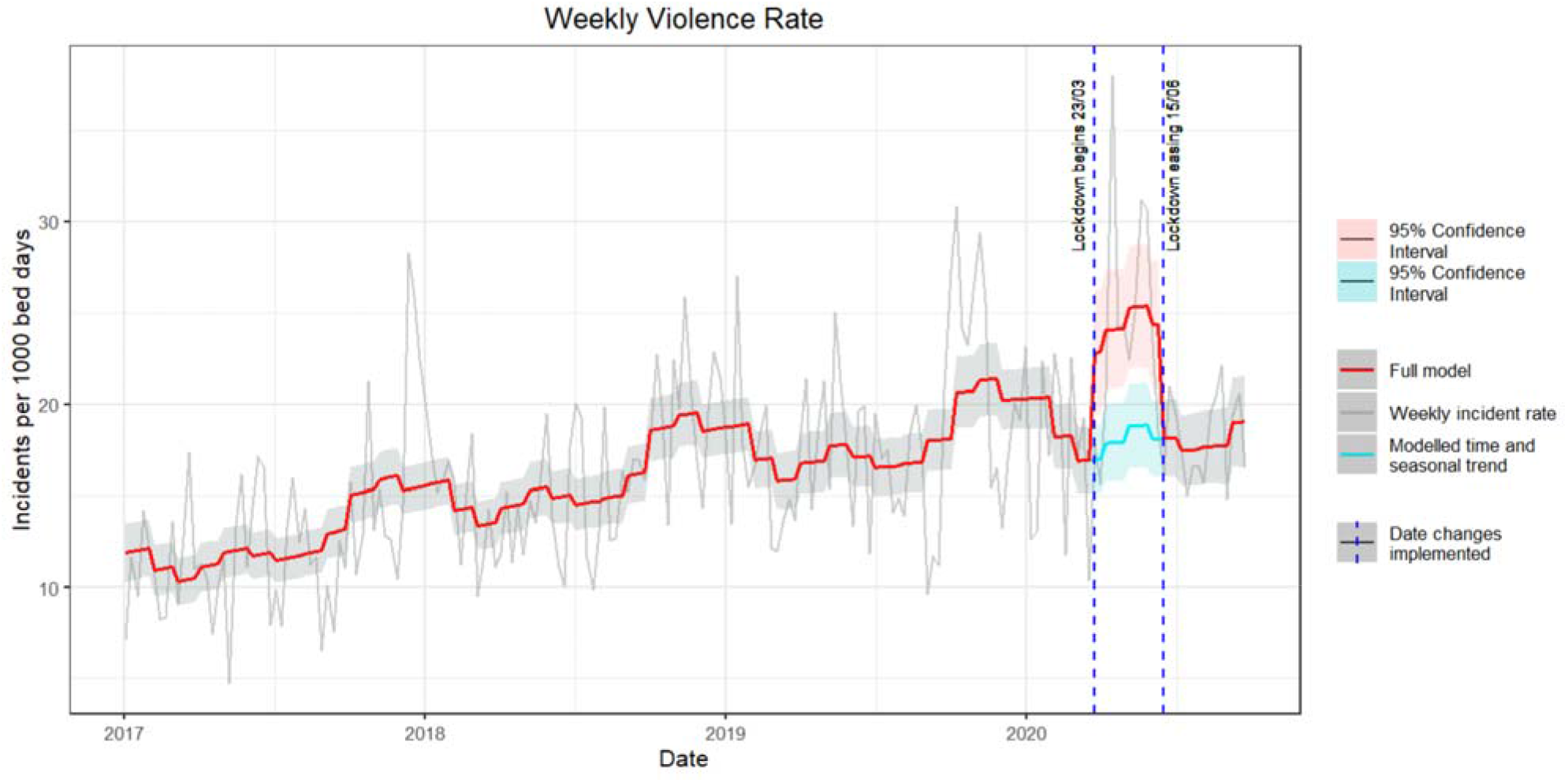
A plot of weekly incidents of violence or aggression per 1000 patient bed days.

Figure 2 part (a) shows the seasonal trend while part (b) shows the time trend. The plots estimate fluctuations around the mean value. The rate of incidents appears to be higher around the end of the year. In addition to this seasonal trend, part (b) suggests there has been an overall increase in the rate of violent and aggressive incidents over the period analysed.

**Figure 2.**
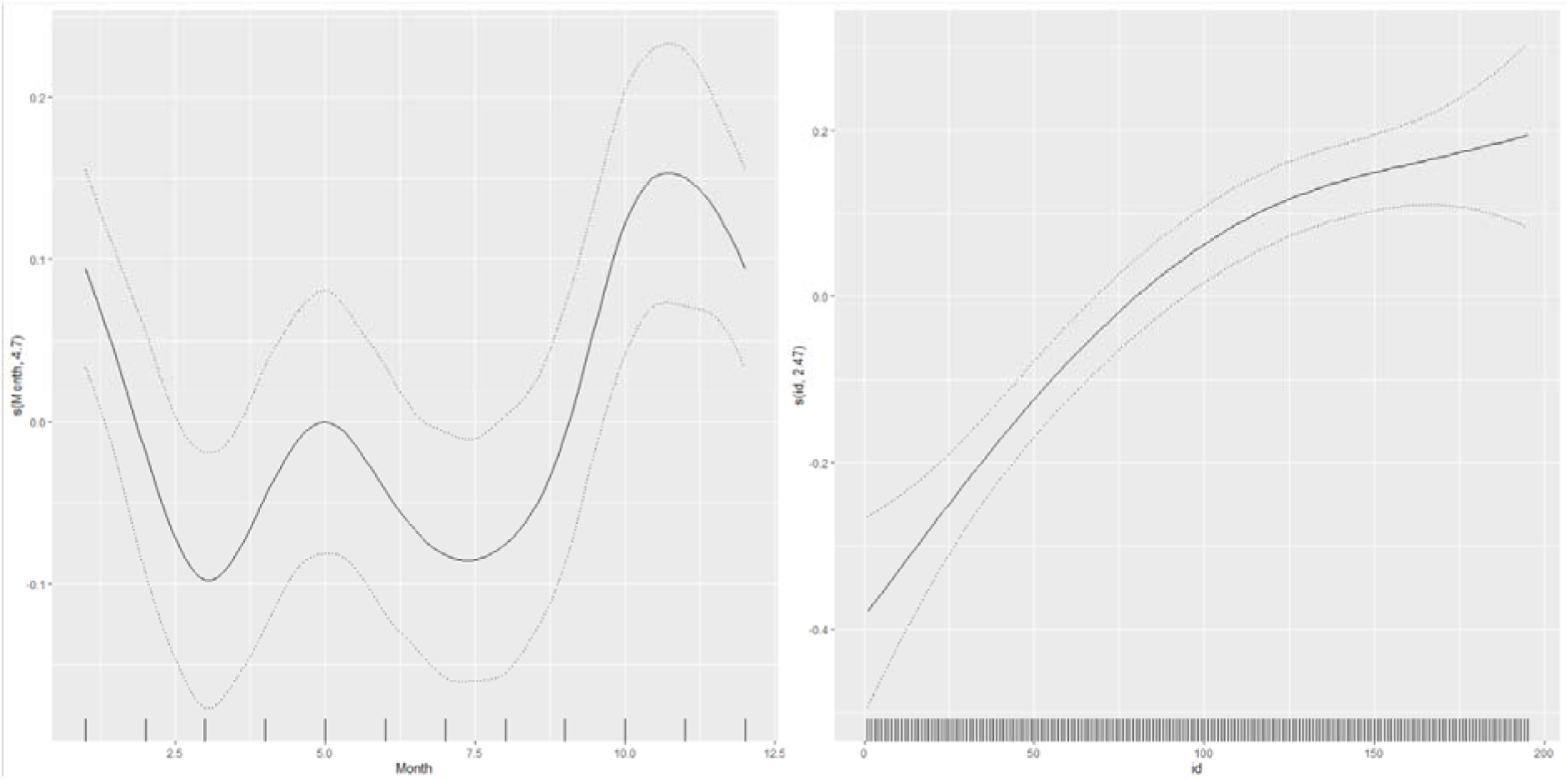
Illustrations of the seasonal and temporal trends identified by the model. The rightmost pane, graph (a), illustrates the seasonal trend while the leftmost pane, graph (b) illustrates the temporal trend.

## Discussion

The present study demonstrates a jump in the rate of violent incidents following the Covid-19 policy changes. The rate of violent incidents relative to the number of occupied bed days jumped by an average of 35% during the period 23^rd^ March to 15^th^ June 2020. There is evidence of an upward trend over the years and a seasonal trend, with the rate of violence peaking towards the end of each year.

Our study has a number of limitations. First of all, there were myriad changes to inpatient life in addition to a likely increase in levels of anxiety due to the threat and uncertainty of the pandemic. There were also likely differences in how strongly the guidelines were applied, given the realities of managing a ward. Our study does not tease these complexities apart to show which factors, if any, increased the rate of violence. Secondly, while we adjusted for seasonal and temporal trends, we did not control for factors pertaining to the composition of the inpatient population that could be associated with the increased rate of violence. A systematic review and meta-analysis found that factors such as a diagnosis of schzophrenia, involuntary admission, a history of alcohol abuse, and a history of violence are associated with higher levels of violence (Lozzino et al., 2015). In response to the pandemic, there was a rapid discharge of psychiatric inpatients, with 2241 more psychiatric inpatients being discharged in March 2020 as compared to February 2020 (Mind, 2020). If inpatients with the lowest levels of risk were discharged during this process, the composition of inpatients would shift towards one exhibiting a higher prevalence of risk factors. This could mean it was not necessarily changes to the ward environment that increased the rate of violence, but rather the fact that more of the remaining inpatients were higher risk and thus the number of incidents relative to occupied bed days would increase.

Despite not delineating underlying mechanisms, our study provides an important description of how the rate of psychiatric inpatient violence increased during the most restrictive phase of Covid-related policies. Our study provides a comprehensive description of violent incidents because it includes all incidents of violence or aggression perpetrated in acute adult wards and PICUs across the Trust. Analysing incidents over almost four years allows us to characterise the underlying time and seasonal trends, and therefore estimate the impact of Covid more precisely given the time of year the policy changes were implemented.

Our findings are relevant in helping to anticipate the effect of future restrictions on inpatient life. Violence on psychiatric wards is a threat to the safety and wellbeing of service users and ward staff alike. This needs to be considered in any cost benefit analysis of future restrictions to the freedoms of psychiatric inpatients. Extra resources may be required to minimise the risk of future increases in the rate of violence resulting from measures to protect physical health.

## Data Availability

We do not have ethnical approval to share the data.

## Acknowledgements

South London and Maudsley NHS Mental Health Trust, Maudsley Hospital, Denmark Hill, Camberwell, SE5 8AZ

## Ethical approval

Ethical approval was received by the South London and Maudsley NHS Trust’s ethical approval committee for clinical audits, service evaluations and other quality improvement projects. The number/ID of the approval is PPF28052020. The project was also approved by the Trust’s Information Governance team.

## Availability of Data and Materials

We do not have ethnical approval to share the data.

